# A versioned, analysis-ready archive of United States State Cancer Profiles county- and state-level estimates

**DOI:** 10.64898/2026.08.24.26361254

**Authors:** Sean Davis

## Abstract

State Cancer Profiles (statecancerprofiles.cancer.gov), maintained by the National Cancer Institute with the Centers for Disease Control and Prevention, is a widely used source of county- and state-level cancer statistics in the United States, used for cancer-center catchment-area surveillance and for geographic studies of cancer burden, screening, and access to care. The site offers no API, no bulk download, and no archive of prior estimates: its sole machine-readable export returns one statistical stratum per HTTP request, and when the underlying data are updated the previous estimates are overwritten and become unrecoverable. This resource provides the complete national county- and state-level extract of all four State Cancer Profiles data topics (incidence, mortality, screening and risk factors, and demographics) as typed, analysis-ready files with the stratifying dimensions as columns, published under pinned, citable version DOIs on Zenodo (concept DOI 10.5281/zenodo.11098814). One version DOI is minted per distinct upstream data *`vintage`*, the set of values the site served between successive replacements. Three vintages have been captured to date; at each observed vintage boundary roughly 97% of estimate values changed, so which vintage an analysis draws on affects its results. From the 2026-08-24 release forward, cells that the upstream site suppresses are retained as typed nulls with an explicit suppression-reason column. Capture has been automated on an approximately monthly cadence since February 2025, and each future upstream revision will be preserved as a new vintage.

**Background & Summary:** State Cancer Profiles (SCP; https://statecancerprofiles.cancer.gov) is a joint National Cancer Institute and Centers for Disease Control and Prevention website that publishes cancer incidence, mortality, screening and risk-factor, and demographic estimates for United States counties, states, and the nation.

## Cancer surveillance, catchment areas, and community outreach

County-level cancer statistics are working inputs to cancer control practice as well as to research. NCI-designated cancer centers carry a formal surveillance obligation: the NCI Cancer Center Support Grant requires each center to define a catchment area, characterize its cancer burden and risk-factor profile, and demonstrate Community Outreach and Engagement (COE) responsive to that burden (J. T. Burus et al. 2023; Spees et al. 2024). Catchment-area surveillance platforms draw on SCP for the county-resolution estimates this work needs (J. T. Burus et al. 2023; Sonawane et al. 2024; Antonio et al. 2024; Munoz et al. 2024; Lowery et al. 2026); one of these, Cancer InFocus, draws directly on SCP and had been licensed by 35 institutions, including 26 NCI-designated centers, as of October 2024 (T. Burus et al. 2025).

Beyond catchment surveillance, published studies use county-level SCP estimates to examine geographic access to oncology care (Shalowitz, Vinograd, and Giuntoli 2015; Y. Zhang et al. 2024; Crowley et al. 2026), environmental and behavioral exposure correlates of cancer burden (Joseph et al. 2022; Wei et al. 2025), community factors associated with screening and mortality (Drake et al. 2025), and incidence in non-metropolitan counties (Jacobson et al. 2026). All of these uses rest on data that can be neither obtained in bulk nor recovered once the site updates: the analysis dataset is whatever SCP served on the day it was queried.

## Bulk access and the vintage problem

As of 23 August 2026, the SCP website provided no public API and no bulk-download facility. Its documentation describes per-table CSV export as the available method for obtaining data, and the site’s sole machine-readable export returns a single statistical *`stratum`* (one cancer site × race/ethnicity × sex × age × stage × area-type combination, covering all areas at once) per HTTP request. Assembling one complete national county- and state-level extract required 15,180 sequential requests (one per published stratum, counted from the deposited scrape catalog after excluding its 3,672 phantom mortality-stage records; see Technical Validation), a workload that rules out casual bulk reuse. The export also encodes withheld cells as asterisks and footnote markers inside otherwise numeric columns, which naive parsing either fails on or silently discards.

County-resolution alternatives do not substitute for this extract. SEER research files require an authenticated data-use agreement and dedicated software (seer.cancer.gov/data/access, accessed 24 August 2026), and the US Cancer Statistics public-use database excludes county-level records under its state data-sharing agreements, reserving them for a restricted-access research data center (cdc.gov/united-states-cancer-statistics, accessed 24 August 2026). To our knowledge, SCP is the only public source of national county-resolution cancer incidence estimates; county cancer mortality is separately queryable through CDC WONDER, but SCP’s derived products (five-year county averages, Joinpoint trend estimates, and modelled screening prevalence) are published nowhere else, and the studies cited above consumed SCP’s numbers as served.

The second limitation is temporal. NCI periodically replaces the estimates (a new SEER/NPCR data submission, revised population denominators, a shifted five-year window), and the site serves only the current values. We refer to each successive set of estimates as a **`vintage`**: the complete set of values the site served during some period, bounded by the upstream replacement. The term follows the real-time-data literature in economics, where a data vintage is a statistical series as it existed on a given date and where the choice of vintage is known to change analytic conclusions (Croushore and Stark 2001, 2003). Cancer surveillance exhibits the same phenomenon: reporting delay and reporting error revise incidence rates for years after first publication, by enough to change the direction of fitted trends (Clegg et al. 2002; Das et al. 2008). At each vintage boundary observed in this archive, approximately 97% of previously published estimate values changed (Technical Validation).

No archive of superseded SCP estimates has existed. The Internet Archive’s Wayback Machine (CDX index, accessed 23 August 2026) holds 35,642 pre-2023 captures of SCP CSV exports, but these collapse to a few hundred distinct strata (under 1.3% coverage of any single vintage, too sparse for reconstruction) while documenting 11 distinct pre-2023 incidence data windows, each overwritten and unretrievable today (`data/wayback_windows.csv`). An analysis built on SCP therefore cannot be reproduced from source after the next upstream refresh, and the estimates that informed a past decision cannot be re-examined.

Prior software does not close either gap. `cancerprof` (WILDS, Fred Hutchinson Cancer Center 2024) wraps SCP’s per-query CSV export in R; it was submitted to rOpenSci software peer review, the review closed without acceptance, and the package was never published on CRAN. It issues one live request per stratum, does not record which vintage the numbers came from, and caches nothing, so repeated runs of the same script can return different values without any indication of change. An earlier R client was abandoned in 2017 (Silent Spring Institute 2017), and a public repository contains an unpublished national SCP sweep feeding a dashboard pipeline, with no release, no registry entry, and no DOI (Community Impact Office, Markey Cancer Center, University of Kentucky 2025). The demand for bulk access is nonetheless visible in the workarounds: at least three CRAN packages ship hand-frozen, single-window SCP slices as example data (Sarkar and Andrews 2025; Pearson 2021; Otto 2025), and the one SCP-derived deposit we located in a generalist data repository is a single-site, single-window analysis slice (Acharjee, Das, and Young 2020).

A live query client and an archive answer different questions: what the site serves today, versus what it served when an analysis was run. The resource described here therefore complements clients such as `cancerprof` rather than competing with them. To our knowledge, no complete SCP extract has previously been published as a citable dataset.

## This resource

This resource addresses both gaps (Figure 1). A scraper enumerates the site’s own query vocabulary, issues one request per stratum, and publishes the complete national county- and state-level extract of all four data topics as typed, analysis-ready CSV and Parquet files in which the stratifying dimensions appear as decoded label columns. From the 2026-08-24 release forward, cells that the site suppresses are retained as typed nulls with an explicit suppression-reason column, so withheld observations remain visible and countable rather than silently absent. Dated captures are published as GitHub releases on an approximately monthly cadence; distinct vintages are identified by content comparison, and each receives a Zenodo version DOI under a single concept DOI (the persistent identifier that always resolves to the newest deposit), following FAIR-oriented dataset-versioning practice (Klump et al. 2021; González-Cebrián et al. 2024). A superseded vintage therefore remains retrievable at its version DOI after the site has replaced it. The design follows published descriptors of re-extracted public and governmental data (Hasell et al. 2020; Xu et al. 2020; Ocagli et al. 2024; Heusden et al. 2025; Karnik et al. 2025; M. Zhang et al. 2025). Three vintages have been captured across 20 releases to date. Vintage boundaries are known only to within the interval between successive captures, and vintages replaced before capture began in 2024 were not archived; the resource is accordingly a prospective record. The deposited vintages are citable and complete independent of continued operation: if capture were to stop, the archive would remain a bounded historical record rather than degrade. Continued capture requires no manual step; the scheduled integration pipeline publishes releases and mints vintage DOIs through the same code path that produced the existing deposits.

**Figure 1:**
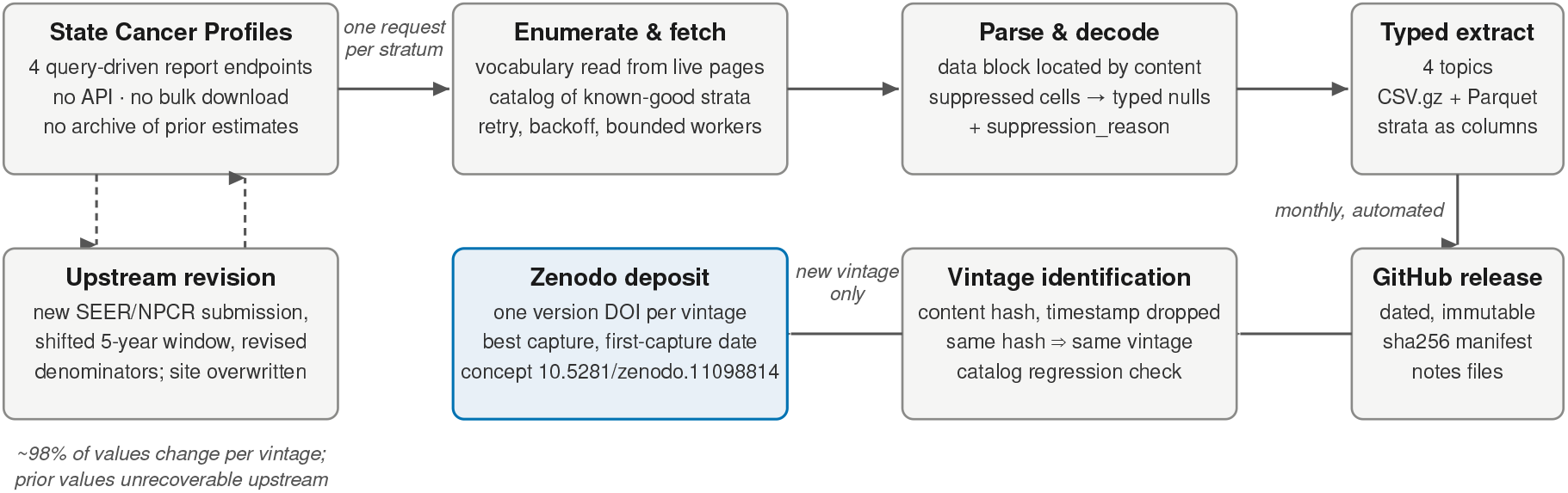
Architecture of the archive. Upper row: monthly capture, one request per statistical stratum against the four State Cancer Profiles report endpoints, with the query vocabulary discovered from the live site, content-based parsing, and suppression decoding into typed nulls with a reason column. Lower row: publication, as immutable dated GitHub releases with checksum manifests; releases are grouped into vintages by content hash, and each new vintage receives a Zenodo version DOI under the data concept DOI. Left: the upstream revision cycle that motivates the archive; estimates are replaced in place and prior values are unrecoverable from the source.

Each of the practice settings above maps to a concrete use of the archive. A catchment-area needs assessment built on a pinned version DOI can be reproduced exactly at the next assessment cycle, and differences from a newer vintage are attributable to upstream revision rather than to a change in the catchment itself. Disparities and access studies can join incidence, mortality, screening prevalence, and demographics at county level in a single query rather than stratum by stratum. And because suppression is recorded rather than silently dropped, a center can identify which populations in its catchment are absent from the public estimates, and methodologists can measure the selection effect of restricting county analyses to unsuppressed areas (Jacobson et al. 2026; Kasheri, Dan, and Nouri 2026).

## Methods

The pipeline converts upstream report pages into typed tables in which every cell of the queried stratum space is accounted for: published values, withheld values of two documented kinds, and combinations the site does not publish are each represented distinctly. From the 2026-08-24 release forward, cells that SCP suppresses are retained as rows with typed null rates and an explicit `suppression_reason` column distinguishing small-count suppression from state-law withholding. We refer to the 2026-08-24 changes (suppression retention, notes preservation, and the Parquet format) as the scraper *`hardening`*. Earlier releases (2024-08-02 through 2026-06-01) predate this behavior and omit suppressed rows entirely.

### Source and extraction unit

SCP serves four query-driven report endpoints: incidence (`incidencerates/`), mortality (`deathrates/`), screening and risk factors (`risk/`), and demographics (`demographics/`). Ap-pending `output=1` to a report URL (the same URL the site’s own “Export Data” link emits) returns the report as CSV. One request fixes one stratum and returns all areas for it; requesting `stateFIPS=00` with `areatype=county` returns every US county plus the national aggregate in a single response, so geography is not an iteration axis. The scraper iterates the cartesian product of the stratifying dimensions per endpoint: cancer site × age × sex × race/ethnicity × stage × area-type for incidence, the same minus stage for mortality (stage is not a mortality dimension upstream; see Technical Validation), topic × risk factor × race/ethnicity × sex × reporting tier for screening and risk factors (the tier, national by state or national by county, is encoded in the state-FIPS query parameter), and area-type × topic × measure × race/ethnicity × sex × age for demographics.

### Query vocabulary and the scrape catalog

The query vocabulary (the valid codes for every dimension) is not hardcoded. It is discovered at run time from the site’s own HTML select elements and JavaScript definition files, so upstream additions (a new cancer site, a new race/ethnicity category) enter the next scrape without a code change. Because most of the cartesian product corresponds to combinations the site does not publish, the scraper maintains a *`scrape catalog`*: an append-only record of every query combination that has ever returned data, with row counts and first/last-seen dates. Monthly runs iterate only the catalog; a full cartesian re-discovery runs quarterly and whenever manually invoked. The catalog doubles as a regression oracle: a combination that returned data at the previous release and returns nothing now is treated as a scrape failure, not as newly dead space, and fails the run. Each release also records the site’s then-current query vocabulary (`select_options.json`) beside the catalog, and a probe warns when upstream adds an identifier the catalog does not know. Requests are issued through a bounded thread pool with exponential backoff on transport errors and server errors; failures are counted and reported, never silently discarded.

### Payload parsing

Each response is a report, not a clean data file: a title block, the data rows, and a footnote block, separated by blank lines. The parser locates the data block by content, scanning for the header line that carries the FIPS or HSA-code field rather than counting fixed line offsets, and reads column names from the header rather than by position. SCP added a 2023-vintage Rural-Urban Continuum Codes column to county reports (first present in this archive at the V1→V2 boundary). The column broke position-based parsing in `cancerprof` (WILDS, Fred Hutchinson Cancer Center 2024) for three of its four topics at county level, a failure we verified against the live site on 2026-08-23 and that the package’s issue tracker has recorded since March 2025; it passed through this parser without incident. Everything outside the data block is preserved verbatim in per-endpoint `notes_<endpoint>.txt` files: the data windows, source registries, suppression-rule text, and data-submission year that constitute the extract’s provenance.

### Suppression decoding

SCP withholds a cell for two documented reasons: counts below 16 in an area-sex-race category (“data has been suppressed to ensure confidentiality and stability of rate estimates”), and state legislation prohibiting release of county-level data. Suppression of this kind is standard practice in federal health statistics (Klein et al. 2002; Parker et al. 2017; Talih et al. 2023), and NCI’s own surveillance program identifies county-level suppression as a material limitation of county cancer reporting (Tatalovich et al. 2022). Adjacent work on census differential privacy likewise shows that upstream disclosure-limitation choices propagate unevenly into small-area health estimates (Santos-Lozada, Howard, and Verdery 2020; Krieger et al. 2021; Li et al. 2023). In SCP’s export the two cases appear as an asterisk or a footnote marker inside otherwise numeric columns. The scraper decodes them before numeric coercion: the row is kept, the rate columns become typed nulls, and `suppression_reason` is set to `suppressed_small_count` or `withheld_state_law`. A marker outside these two would surface as a null rate with no reason code; Technical Validation shows the deposited file contains none. A user can therefore distinguish “suppressed for small counts”, “legally withheld”, and “combination never published”: three states that are indistinguishable in an extract that drops non-numeric rows, as this scraper’s own releases did before 2026-08-24.

### Vintage identification

Release artifacts are assigned to vintages by content, not by date: each topic file is hashed (SHA-256) after dropping the single scrape-time-varying column (`_extracted_at`), and releases sharing content hashes captured the same upstream data. Vintage identity is keyed on the registry-observed incidence and mortality topics only; the screening/risk-factor and demographics topics are modelled or survey products that refresh on their own cadence (Usage Notes). Candidate boundaries are confirmed by decomposing each between-release difference into new query strata versus changed values on strata common to both releases: a scope expansion of the scraper adds strata but changes no common values, whereas an upstream revision changes nearly all of them (Technical Validation). When the scraper’s own output format changes, as at the 2026-08-24 hardening, content hashes no longer compare across the format change, and vintage continuity is instead established by a value-level comparison on common strata.

### Harmonized view

Alongside the immutable per-release files, the pipeline derives a harmonized cross-vintage view. Harmonization decisions (label crosswalks, column renames, the reclassification of county rows mislabelled by an upstream quirk) are recorded as data in a checked-in crosswalk file, not hardcoded, and the transformation enforces strict accounting: harmonized rows plus dropped rows must equal original rows, and any imbalance fails the build. Published release bytes are never regenerated or edited; corrections are new releases.

### Archiving and deposit

A scheduled GitHub Actions workflow scrapes monthly, validates against the catalog, writes a manifest of SHA-256 checksums, row counts, and byte sizes for every artifact, and publishes a dated GitHub release. When the manifest identifies a new vintage, the same code path mints a new version DOI on the data concept. Each vintage’s deposit carries the *`best`* capture of the vintage (the most complete release, which is not always the first), sets the publication date to the *`first`* capture, and lists every GitHub release tag that captured the vintage in its related identifiers. Code and data are archived under separate Zenodo concepts; the data concept (10.5281/zenodo.11098814) is the citable identity of this resource.

## Data Records

The archive is deposited on Zenodo under concept DOI 10.5281/zenodo.11098814, which always resolves to the newest vintage. Each distinct vintage carries its own version DOI (Table 1).

**Table 1:** Data vintages, their Zenodo version DOIs, and the GitHub releases that captured them. The publication date of each version DOI is the date of the vintage’s first capture; the deposited bytes are its most complete capture.

| Vintage | DOI | Releases capturing it | First captured | Best capture | Topics deposited |
| --- | --- | --- | --- | --- | --- |
| V1 | [10.5281/zenodo.12685787](https://doi.org/10.5281/zenodo.12685787) | 1 | 2024-08-02 | 2024-08-02-1 | incidence, mortality |
| V2 | [10.5281/zenodo.22085047](https://doi.org/10.5281/zenodo.22085047) | 13 | 2025-02-10 | 2026-02-01 | incidence, mortality |
| V3 | [10.5281/zenodo.22085273](https://doi.org/10.5281/zenodo.22085273) | 6 | 2026-04-01 | 2026-08-24 | all four |

Each vintage deposit contains one gzipped CSV per data topic captured: the two registry topics, incidence and mortality, for V1 and V2, and all four topics from V3 (Table 1). The V3 deposit (best capture `2026-08-24`, the first from the hardened scraper) adds a Parquet file per topic, the per-endpoint notes files, the scrape catalog, the query vocabulary (`select_options.json`), a manifest with per-file SHA-256 checksums, and a rendered README with citation guidance. Earlier vintages are CSV-only captures made before the hardening and are preserved byte-for-byte as scraped; under the immutability rule they will not gain Parquet or suppression columns retroactively. The `2026-08-24` release comprises 20,275,578 rows across the four topics (incidence 11,048,853, mortality 5,866,632, demographics 3,281,295, screening and risk factors 78,798), totalling approximately 383 MB as gzipped CSV or 132 MB as Parquet.

All four topic tables share a common shape, one row per (stratum × area), and one geographic universe: 3,193 distinct five-digit county-level area codes per topic (United States counties, the District of Columbia, and Puerto Rico municipios, plus a small number of upstream aggregate rows carrying nonstandard codes, preserved as served), together with state and national tiers. The incidence, mortality, and demographics tables record the query tier in `areatype`; the upstream risk report carries no area-type field, so the risk table records its tier in `statefips_query` (`00`, national by state; `99`, national by county). Table 2 lists the columns of the incidence table, the widest of the four, as read from the deposited Parquet file. Mortality omits `percent_of_cases_with_late_stage` and its `stage` column is constant; demographics and screening/risk-factor tables substitute their own measure columns (population characteristics and modelled prevalence estimates with confidence intervals, respectively) and their own topic/measure dimension columns, but carry the same geographic keys, provenance columns, and decoded stratifier labels.

**Table 2:**
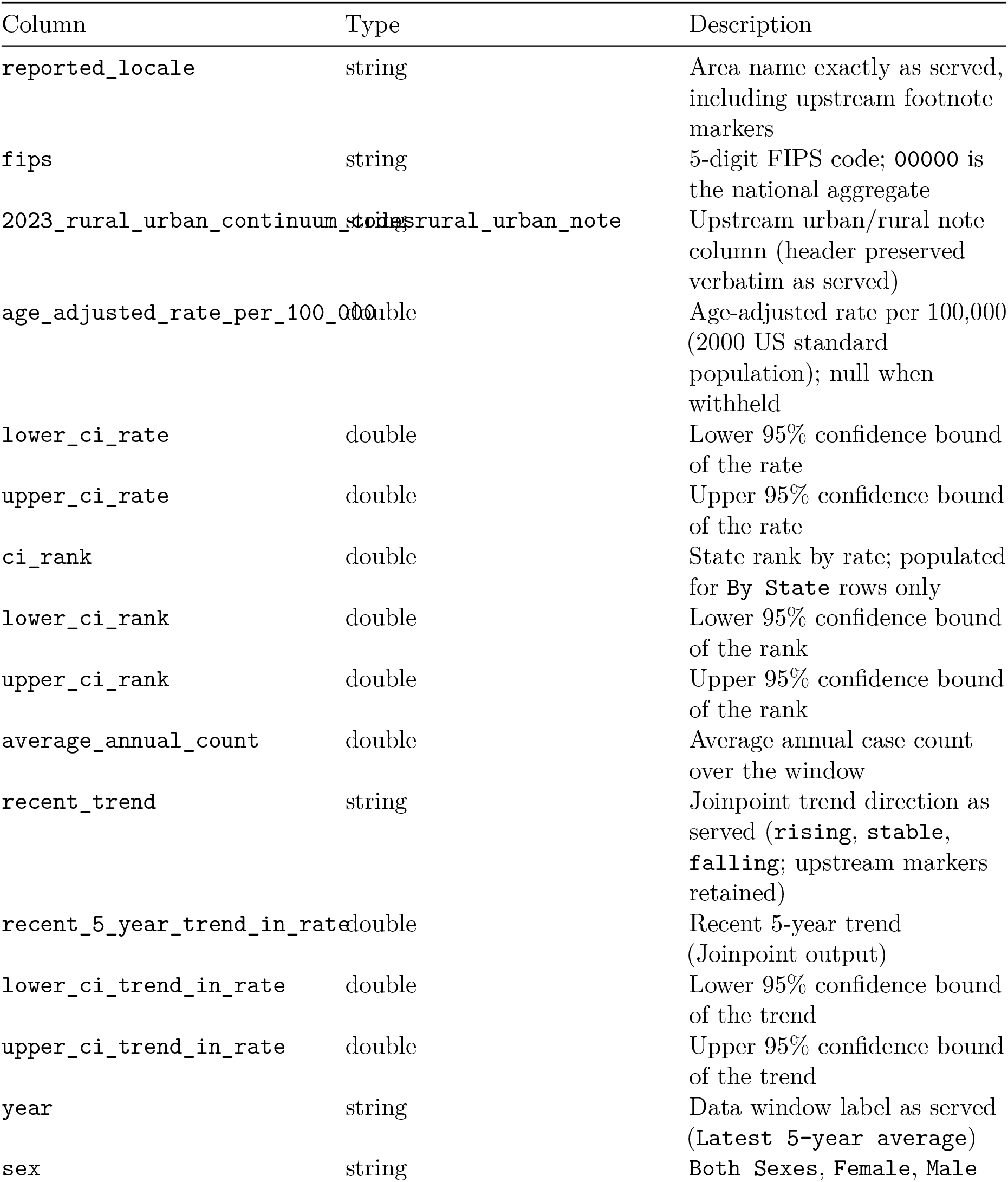
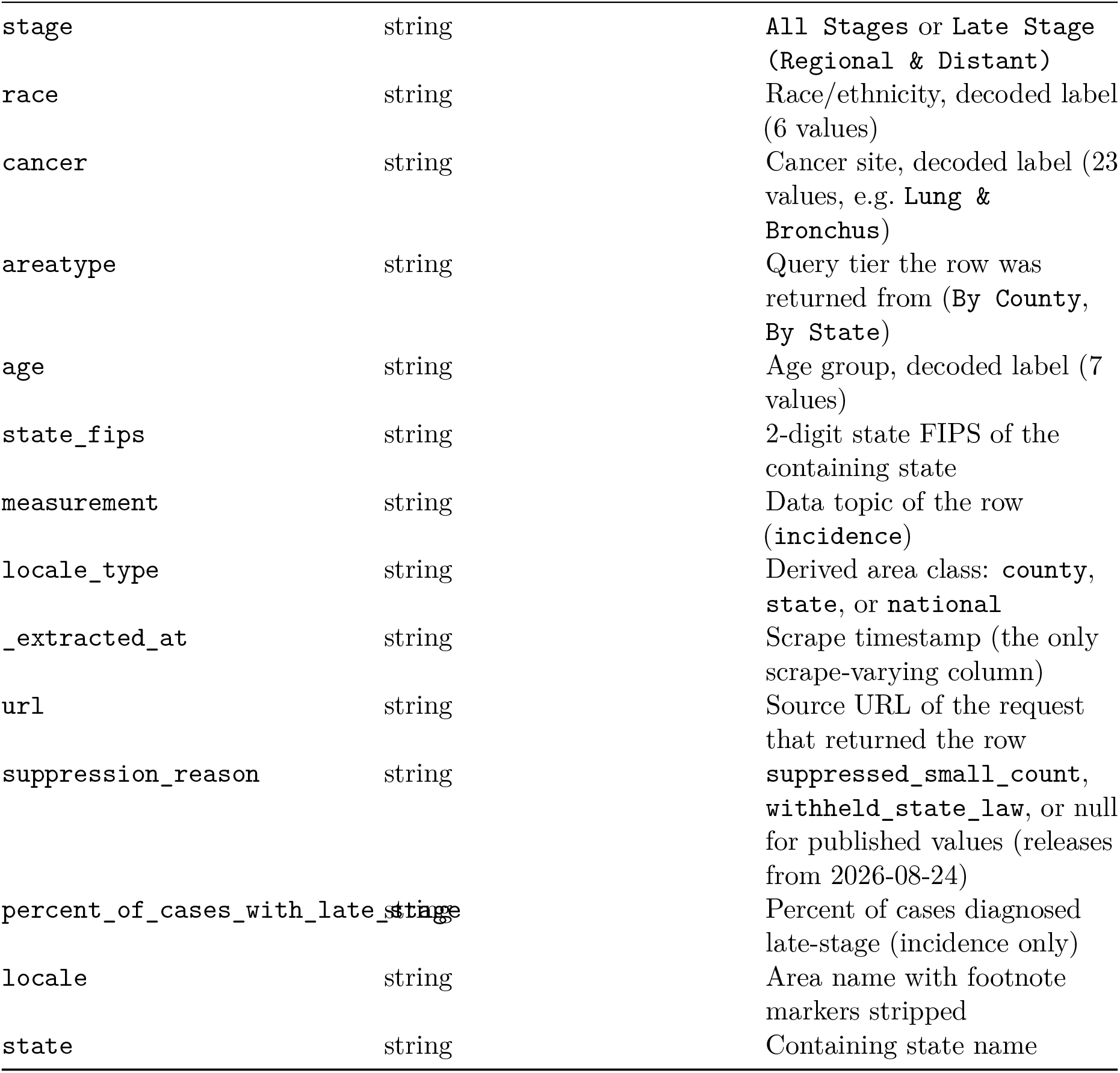
Column schema of the incidence table, read from the deposited 2026-08-24 Parquet artifact (`data/schema_incidence.csv` is derived from the deposit by `scripts/derive_figure_data.py`).

Figure 2 illustrates the county-level content of a single vintage: the age-adjusted all-sites incidence rate, and the cell status of one finer stratum in which suppression becomes visible.

**Figure 2:**
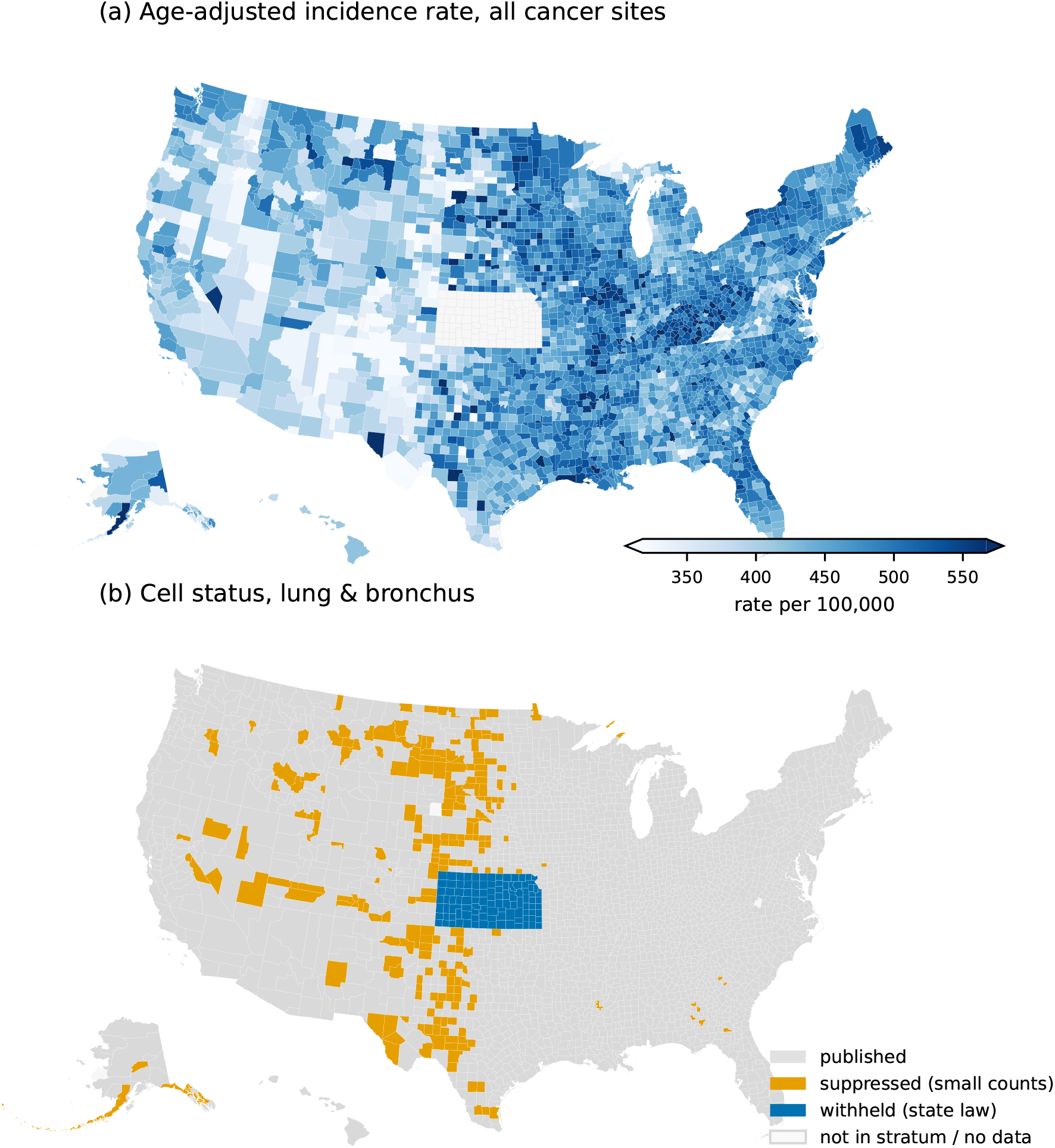
County-level content of vintage V3 (release 2026-08-24). (a) Age-adjusted incidence rate per 100,000, all cancer sites, all races, both sexes. Kansas publishes no county-level incidence in any vintage and appears as missing. (b) Cell status for one finer stratum (lung and bronchus, all races, both sexes): published rate, suppressed for small counts, or withheld under state law (all Kansas counties); counties absent from the stratum are shown as no data. The color scale in (a) is truncated at the 2nd and 98th percentiles. Puerto Rico is omitted from the maps; boundaries are 2010 Census counties, matching the legacy county codes the site serves.

The complete capture history (every dated release, including the several releases that captured each vintage) is public on GitHub (https://github.com/seandavi/state-cancer-profile-scraper/releases) with per-release manifests in the repository. The harmonized cross-vintage view is a derived artifact, regenerated deterministically from the release files by the pipeline (`scps.normalize`); it is not separately deposited. Data are licensed CC-BY-4.0; the scraper and pipeline code are MIT-licensed. SCP’s published data-use terms restrict use to statistical reporting and analysis and prohibit attempts to identify any person or establishment (statecancerprofiles.cancer.gov/dataUseRestrictions.html, accessed 24 August 2026); the archive redistributes only the aggregate, already-suppressed statistics the site publishes, and those terms apply unchanged to its contents. As publications of United States federal agencies, the underlying estimates are not subject to domestic copyright, so their redistribution requires no license from the producing agencies; the carried-forward terms constrain use, not redistribution. The CC-BY-4.0 license covers the archive’s compilation, documentation, and derived artifacts.

The archive satisfies the FAIR principles (Wilkinson et al. 2016): each vintage is findable and citable through its version DOI under a stable concept DOI; all artifacts are accessible over plain HTTPS without registration, under CC-BY-4.0; the typed Parquet and CSV files, keyed on FIPS codes, are interoperable with Census and TIGER products and with any columnar toolchain; and reuse is supported by per-file checksums, per-release provenance (source URLs, extraction timestamps, notes files), and immutable release bytes. The long-format typed tables are likewise suitable for direct ingestion into statistical and machine-learning pipelines: the stratifying dimensions are ordinary columns, the suppression-reason field gives missingness an explicit semantics rather than silent absence, and a pinned version DOI fixes training data exactly.

The archive is additionally mirrored to a public Hugging Face dataset repository (https://huggingface.co/datasets/seandavis/state-cancer-profiles). The mirror is byte-identical to the deposits (every mirrored file’s SHA-256 is verified against the release manifest before commit), each vintage is a tagged commit named for the Zenodo version DOI it mirrors (`zenodo-v1` through `zenodo-v3`), and the dataset card directs citation to the concept DOI; no DOI is minted on Hugging Face, and Zenodo remains the archival copy. The mirror exists for direct query access. DuckDB resolves `hf://` paths against the repository without authentication, so the current vintage can be evaluated in one statement:

~~~
**SELECT** count(*)
**FROM** ‘hf://datasets/seandavis/state-cancer-profiles/state_cancer_profiles_incidence.parquet’
~~~

which returns the incidence row count recorded in the release manifest (11,048,853; verified live in Technical Validation). Because the mirror’s hosting serves cross-origin and HTTP range requests, the same queries also run in a browser through DuckDB-WASM with no installed software; the Zenodo deposits themselves do not serve cross-origin requests, so the in-browser path applies to the mirror only.

### Technical Validation

#### Release integrity

Every deposited artifact is listed in a checked-in manifest with SHA-256 checksum, byte size, and row count, and the Zenodo deposits were verified byte-identical against these manifests at publication. Table 3 reports per-release, per-topic row counts and content checksums for the full capture history.

**Table 3:** Per-release, per-topic row counts and content checksums (SHA-256 of the file after dropping the scrape-timestamp column) for all releases. Identical checksums within a column identify releases that captured the same vintage. Mortality row counts in releases before 2026-08-24 include the phantom-stage duplication described in the text.

| Release | Topic | Rows | Content sha256 (first 12) |
| --- | --- | --- | --- |
| 2024-08-02-1 | incidence | 934,287 | 95bb771c6742 |
| 2024-08-02-1 | mortality | 774,770 | 1f40af102eb8 |
| 2025-02-10 | incidence | 1,014,762 | 2ae50c264b32 |
| 2025-02-10 | mortality | 782,632 | 7e766d8f9630 |
| 2025-03-01 | incidence | 1,014,762 | 2ae50c264b32 |
| 2025-03-01 | mortality | 782,632 | 7e766d8f9630 |
| 2025-04-01 | incidence | 1,014,762 | 2ae50c264b32 |
| 2025-04-01 | mortality | 782,632 | 7e766d8f9630 |
| 2025-05-01 | incidence | 1,013,440 | 99a237b478d8 |
| 2025-05-01 | mortality | 782,632 | 7e766d8f9630 |
| 2025-06-01 | incidence | 1,014,762 | 2ae50c264b32 |
| 2025-06-01 | mortality | 782,632 | 7e766d8f9630 |
| 2025-07-01 | incidence | 1,014,762 | 2ae50c264b32 |
| 2025-07-01 | mortality | 782,632 | 7e766d8f9630 |
| 2025-08-01 | incidence | 1,014,762 | 2ae50c264b32 |
| 2025-08-01 | mortality | 782,632 | 7e766d8f9630 |
| 2025-09-01 | incidence | 1,014,762 | 2ae50c264b32 |
| 2025-09-01 | mortality | 782,628 | 342cb495c3f1 |
| 2025-10-01 | incidence | 1,014,762 | 2ae50c264b32 |
| 2025-10-01 | mortality | 782,632 | 7e766d8f9630 |
| 2025-11-01 | incidence | 1,014,762 | 2ae50c264b32 |
| 2025-11-01 | mortality | 782,632 | 7e766d8f9630 |
| 2025-12-01 | incidence | 1,014,762 | 2ae50c264b32 |
| 2025-12-01 | mortality | 782,632 | 7e766d8f9630 |
| 2026-01-01 | incidence | 1,014,762 | 2ae50c264b32 |
| 2026-01-01 | mortality | 782,632 | 7e766d8f9630 |
| 2026-02-01 | incidence | 1,014,762 | 2ae50c264b32 |
| 2026-02-01 | mortality | 782,632 | 7e766d8f9630 |
| 2026-04-01 | incidence | 1,354,812 | 5e7f9f50ef76 |
| 2026-04-01 | mortality | 928,958 | 2ecc04fa1975 |
| 2026-05-01 | incidence | 1,354,812 | 5e7f9f50ef76 |
| 2026-05-01 | mortality | 928,958 | 2ecc04fa1975 |
| 2026-05-27 | incidence | 1,354,812 | 5e7f9f50ef76 |
| 2026-05-27 | mortality | 928,958 | 2ecc04fa1975 |
| 2026-05-28 | demographics | 3,310,984 | 93305466c13a |
| 2026-05-28 | incidence | 1,476,853 | db2b0672dfdf |
| 2026-05-28 | mortality | 1,034,042 | ec50eace83b0 |
| 2026-05-28 | risk | 83,429 | 663798d89d33 |
| 2026-06-01 | demographics | 3,310,984 | 6a8dfedb0c07 |
| 2026-06-01 | incidence | 1,476,853 | db2b0672dfdf |
| 2026-06-01 | mortality | 1,034,042 | ec50eace83b0 |
| 2026-06-01 | risk | 83,429 | 36fc0cc9a8a9 |
| 2026-08-24 | demographics | 3,281,295 | 5b37483e2c2a |
| 2026-08-24 | incidence | 11,048,853 | a120cd6e0f73 |
| 2026-08-24 | mortality | 5,866,632 | 16f9f3970739 |
| 2026-08-24 | risk | 78,798 | 6a370ab842e9 |

Two historical releases are defective partial scrapes rather than distinct vintages: `2025-05-01` incidence is missing exactly one query stratum (1,322 rows) relative to its neighbours, and `2025-09-01` mortality is missing 4 rows; both revert identically at the next release, which upstream revisions do not do. Both are recorded as defective in the vintage assignments and excluded from best-capture selection.

In releases before `2026-08-24` the scraper iterated a stage dimension that the mortality endpoint ignores, so historical mortality tables contain each row twice: once labelled all-stages and once spuriously labelled late-stage. Row counts for those releases must be halved after deduplication, and the phantom late-stage labels discarded; the hardened scraper no longer iterates stage for mortality.

#### Vintage assignment

Within each vintage, all clean captures are byte-identical after dropping the extraction timestamp (eleven clean V2 releases share one content hash per topic), so the release-to-vintage grouping is not sensitive to capture date. At the two observed vintage boundaries, decomposition into new versus common strata shows the signature of upstream revision (comparing age-adjusted rate values on strata common to both releases, computed from the deposited files by the derivation script): from V1 to V2, 97.4% of incidence values changed across 902,787 common rows (97.4% of mortality on 733,172); from V2 to V3, 97.3% changed on 981,550 published-in-both rows (96.1% of mortality on 376,745 after collapsing the phantom stage dimension). By contrast, the within-V3 scraper scope expansion of `2026-05-28` added 122,041 incidence rows while changing zero values on 1,299,615 common rows, and the `2026-08-24` hardening changed zero values on 1.99 million common incidence and mortality rows; both are therefore capture-scope changes, not new vintages.

The per-county comparison in Figure 3 is computed from the deposited artifacts: of 2,932 counties with a published all-sites rate in both vintages, 99.7% have different values, with a median absolute difference of 9.7 per 100,000 (interquartile range 4.4–18.6). Near-total value change is expected from the window advance alone, since consecutive vintages share four of five years of a moving average; the fraction changed establishes only that revision is pervasive.

**Figure 3:**
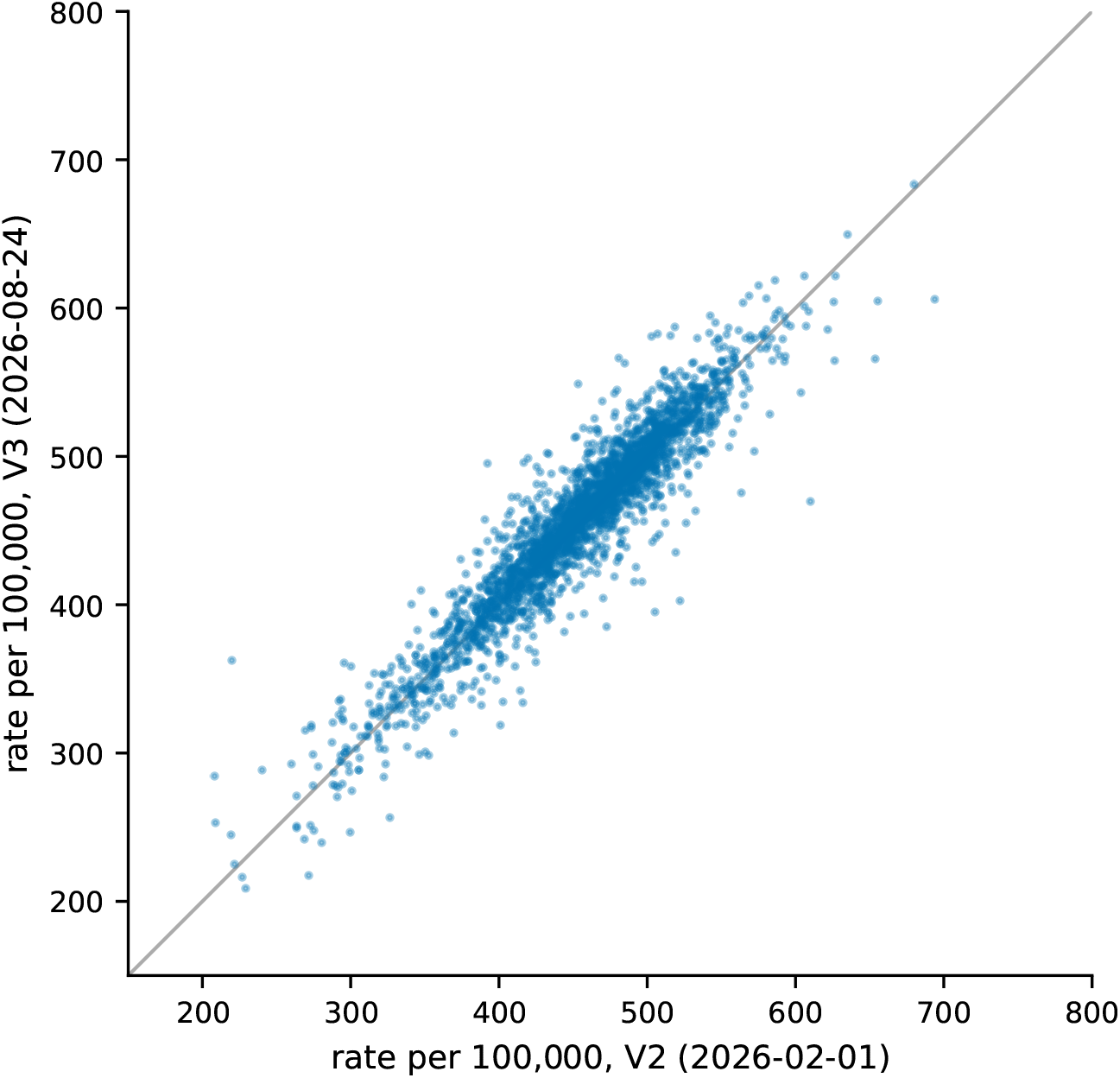
County-level age-adjusted all-sites incidence rates in vintage V2 (release 2026-02-01) against the same counties in vintage V3 (release 2026-08-24); the diagonal is identity. Nearly every county’s published rate changed at the vintage boundary. Axes are truncated at 800 per 100,000; one county lies beyond both limits (Union County, Florida, FIPS 12125: 1,248 in V2, 1,215 in V3).

The magnitude distribution, together with the reporting-delay literature cited in Background, is the evidence that vintage choice matters, and the non-independence of consecutive vintages is restated as a usage caution in Usage Notes.

Vintage boundaries are bracketed, not dated: V1→V2 is bounded by a six-month gap in captures and V2→V3 by a missing monthly release. An upstream vintage could in principle have come and gone inside either window uncaptured; the archive claims only the vintages it holds.

#### Coverage

Mortality coverage is essentially complete and stable (3,082–3,087 distinct published counties across the three vintages’ best captures, no state ever absent), consistent with its vital-statistics origin. Incidence coverage depends on state registry publication decisions and grows across vintages: 2,694 counties in V1, 2,933 in V2, 3,029 in V3 (computed from the deposited best captures). Kansas has 0 published county incidence cells in any vintage (state-level Kansas incidence appears only from `2026-05-28`), while its county-level mortality covers 104 of Kansas’s 105 counties in V3; the gap is the missing block in Figure 2 (a). Indiana county incidence is absent in V1 and V2 and present, all 92 counties, from V3. Minnesota, Nevada, and Virginia county incidence are missing only in V1.

#### Schema stability

Across all three vintages the category vocabularies are identical (23 cancer sites, 6 race/ethnicity groups, 3 sex values, 7 age groups, 2 stage values), and exactly one column was ever added upstream (the 2023 Rural-Urban Continuum Codes note column, at the V1→V2 boundary). The drift audit behind this finding compared column lists, inferred types, and full category vocabularies across every release.

#### Suppression census

In post-hardening files suppression is countable directly from `suppression_reason`. Across the full V3 incidence file, 9,369,770 cells are suppressed for small counts and 202,230 are withheld under state law, the latter entirely in Kansas; the per-state census is `data/suppression_census.csv`. Published, suppressed, and withheld cells sum to the file’s 11,048,853 rows, so no unrecognized marker produced a null rate without a reason code. In the stratum mapped in Figure 2 (b), lung and bronchus, all races, both sexes, the site returned 3,143 county cells: 2,797 published, 241 suppressed for small counts, and 105 (every Kansas county) withheld under state law, matching a reconciliation of the same stratum pulled live from the site’s export URL on 2026-08-23. Historical releases dropped withheld rows, which is why post-hardening captures of the same vintage are substantially larger than earlier ones: the difference consists of retained suppressed cells and the added state-level tier, not changed estimates.

#### Mirror integrity

Every file on the Hugging Face mirror was SHA-256-verified against the release manifest before commit. As an end-to-end check, this document queries the live mirror at render time: a `count(*)` over the mirrored incidence Parquet returns 11,048,853 rows, equal to the row count in the deposited manifest, and rendering fails on any mismatch.

#### Cross-topic joins

The four topics share FIPS geographic keys. Joining the all-sites county strata of the V3 deposit on `fips` (computed by the derivation script) matches 2,980 counties from 3,029 incidence and 3,084 mortality counties; the 104 mortality-only counties are the Kansas asymmetry described above. The demographics topic carries 10 distinct race/ethnicity labels, of which 5 map exactly to the incidence/mortality vocabulary through the checked-in crosswalk; the remainder name populations with no exact equivalent and stay topic-local rather than being forced into a lossy alignment.

### Usage Notes

#### Vintages are not independent observations

Incidence and mortality estimates are five-year averages, and consecutive vintages share four of their five years. Differencing two vintages does not estimate change over time; it largely re-observes the same person-years after revision. Users who need temporal contrasts should use the year windows documented in the notes files, not vintage-over-vintage differences. Notes files exist only from the `2026-08-24` release: the earlier scraper discarded the report notes, so for V1 and V2 the five-year window underlying the estimates is not recoverable from the deposits; their `year` column records only `Latest 5-year average`.

#### Treatment of 2020

SCP displays 2020 incidence but excludes it from its trend fits owing to pandemic-era reporting disruption. Downstream trend work must handle 2020 explicitly rather than inherit its inclusion.

#### Trend statistics are software outputs

The trend columns are Joinpoint model outputs (Kim et al. 2000; Clegg et al. 2009), and Joinpoint’s defaults have changed materially over its lifetime (Kim et al. 2022, 2023). Average annual percent change values should not be compared across vintages without confirming the producing software version in the notes files.

#### Coverage asymmetries

Joining incidence to mortality on FIPS silently drops all Kansas counties from the incidence side (Kansas mortality is complete; Kansas county incidence does not exist in any vintage). Indiana county incidence exists only from V3, so any cross-vintage comparison involving Indiana compares presence against absence. See Technical Validation for the full list of V1 gaps.

#### Filter on areatype, not locale_type, in pre-2026-05-28 files

In those releases `locale_type` misclassifies tens of thousands of county rows (Louisiana parishes, Alaska boroughs, independent cities, and DC) as `other`. The risk table has no `areatype` at all; filter its tier on `statefips_query` instead. The harmonized view corrects the misclassification; the immutable release bytes do not.

#### Suppression semantics differ by era

In releases from `2026-08-24` forward, suppressed cells are present as rows with typed nulls and a reason code. In all earlier releases, suppressed cells are simply absent, and suppression there is recoverable only by differencing against an expected cross-product. The archive does not un-suppress anything; it makes suppression countable. Published small-area studies restrict their samples to unsuppressed areas without being able to characterize what was withheld (Jacobson et al. 2026; Ladas and Towery 2026; Kasheri, Dan, and Nouri 2026).

#### Screening and risk-factor estimates are modelled

They are small-area model estimates combining BRFSS and NHIS survey data (Raghunathan et al. 2007; Liu et al. 2019, 2025), not registry-observed counts. (Related county-level modelled prevalence products exist, for example CDC PLACES; the estimates here are SCP’s own product as served.) They can change between captures because the model was refit, not because the underlying population changed. For this reason vintage identity is keyed on incidence and mortality only; the Zenodo deposit for a vintage carries the newest screening/demographics edition captured during that vintage, and earlier within-vintage editions remain available, hash-verified, in the GitHub releases.

## Code availability

The scraper, pipeline, validation, and deposit code are available at https://github.com/seandavi/state-cancer-profile-scraper under the MIT license, and are archived per-release on Zenodo under a separate software concept (10.5281/zenodo.13174526). Every data release records the exact repository commit that produced it (`gh_hash.txt`), so any capture can be traced to the code that made it. The figure and table inputs in this article are derived from the deposited artifacts by `manuscript/scripts/derive_figure_data.py` in the same repository.

## Data availability

All data are openly available under CC-BY-4.0. The canonical citable deposit is Zenodo concept DOI 10.5281/zenodo.11098814, with one version DOI per vintage (Table 1). The complete dated capture history is available as GitHub releases at https://github.com/seandavi/state-cancer-profile-scraper/releases, and a byte-verified mirror for direct `hf://` query access is maintained at https://huggingface.co/datasets/seandavis/state-cancer-profiles; the mirror carries no DOI, and citation should use the Zenodo DOIs.

## Ethics statement

This work uses only publicly available, aggregate statistics published by the National Cancer Institute and the Centers for Disease Control and Prevention. No individual-level data were accessed and no human-subjects research was conducted; institutional review board approval was not required.

## AI use disclosure

AI coding and writing assistants (Anthropic Claude) were used in this work: to write and revise pipeline and validation code, to run the audits summarized in Technical Validation, and to edit manuscript text. All AI-produced code and text were reviewed, edited, and approved by the author, who takes full responsibility for the content. A working log of AI sessions and the disposition of their output (accepted, modified, or rejected) is kept in the project repository.

